# Development and validation of risk prediction models for childhood, teenage and young adult cancers: research protocol and statistical analysis plan

**DOI:** 10.1101/2024.06.25.24309467

**Authors:** Defne Saatci, Anthony Harnden, Julia Hippisley-Cox

## Abstract

**Background:** Childhood, teenage and young adult (CTYA, 0-24 years) cancers are rare and diverse, making timely diagnosis challenging. Studies based on adult cancers suggest that the development and integration of clinical decision tools in primary care aid earlier cancer detection, yet, these have not been explored for CTYA cancers.

**Aim:** To develop and validate a primary care-based risk prediction tool to identify CTYA who are at increased risk of cancer.

**Methods and analysis:** Using the QResearch Database, a nationally representative primary care database, we will generate an open cohort of children, teenagers and young adults (0-24 years) who were registered with a GP between 1^st^ January 1998 and 31^st^ December 2019. CTYA will be followed up from the date at which the first cancer-relevant symptom is recorded in the records (index date) until the date of cancer diagnosis/6-months, whichever comes first. Candidate variables will include symptoms, signs, blood test results and demographic factors. Model derivation will include two approaches, Cox regression and logistic regression. Apparent performance of the derived model will be explored and subsequently internally-externally cross-validated to investigate performance heterogeneity and geographical transportability.

## Introduction

Childhood and teenage cancer ranks as the 6^th^ leading cause of total cancer burden worldwide and is associated with significant long-term morbidity^1^. Cancer is the commonest cause of mortality by disease among children and young people in the United Kingdom (UK)^2^. Delayed cancer detection is a known contributing factor^3^, with presentation at advanced stages recognised to reduce survival^4^. The UK has longer time-to-diagnosis across childhood cancers compared to other high-income countries^5,6^, as well as higher mortality rates across teenage and young adult cancers^7^. This is an ongoing concern for young people with cancer, who, in a recent national survey, highlighted early diagnosis research as one of their top ten research priorities^8^. This collectively highlights a clear health challenge and there is a pressing need to improve early detection of these cancers in the UK. Indeed, this is in line with the National Health Service (NHS) Long Term Plan (2019)^9^, UK Cancer Reform Strategy (2015)^10^ and Childhood Cancer and Leukaemia Group (CCLG) Strategic Plan (2020)^11^.

The non-specific presentation and relative rarity of childhood, adolescent and young adult cancers (CTYA) pose difficult diagnostic challenges to clinicians^12^ and increase the possibility of delays. This is particularly relevant to general practitioners (GPs) who encounter CTYA cancer patients at the earliest stages of the disease. National awareness initiatives, such as HEADSMART^5^, have been employed to address this challenge in the UK and although this initiative contributed to substantial improvements in diagnostic intervals in central nervous system (CNS) tumours, the national time-to-diagnosis target of 4 weeks has not been reached for all age groups^13^. Furthermore, GPs remain unconfident in diagnosing childhood cancers even after taking part in this initiative^14^. Similarly, recent findings of the “Accelerate, Coordinate, Evaluate” (ACE) programme demonstrated ongoing delays in referrals and cancer diagnosis in TYA^15^. Clearly, novel approaches need to be explored to supplement current pathways.

Computer-based clinical decision tools (CDTs) are increasingly being used in clinical settings, supporting medical decision-making where challenges such as diagnostic uncertainty are present^16^. Overall, CDTs have been reported to reduce diagnostic errors^17^, improve clinical practice and patient care^17^. In primary care settings, recent evidence suggests that technology-based CDTs provide the most successful interventions in reducing diagnostic inaccuracies^18^.The potential for CDTs in cancer diagnosis have been highlighted as an “area of extraordinary opportunity”, with promising developments seen in several adult cancers^19^. A recent systematic review^20^ has shown that these tools for cancer risk prediction have the potential to improve decision-making and clinical service outcomes, as well as one study showing reduction in time-to-diagnosis. Despite these advancements in adult cancers, however, CDTs have not been explored in CTYA cancers.

Accordingly, in this study, we plan to use QResearch Database, one of the largest GP electronic health record database in the UK, to explore ways to detect childhood and TYA cancers earlier by developing a novel GP-based risk prediction tool for CTYA cancers.

## Methods and Analysis

### Data sources and Study Population

#### Data Sources

QResearch Database is a nationally representative primary care database consisting of over 35 million anonymised health records from approximately 1300 general practices (GPs) in England (∼ 20% UK population)^21,22^. Records consist of patient-level demographic information (i.e., year-of-birth, sex, self-assigned ethnicity), as well as clinical information, including cancer diagnoses and clinical presentations. Primary care records are linked to hospital admission, civil registration and the National Cancer Registry data, where linkage is based on an individual patient’s anonymized NHS number. This number is valid and complete in 99.8% of primary care/civil registry data and 98% of hospital admissions data^22^.

#### Study Population

Model development and validation will use an open cohort of children, teenagers and young adults (from birth up to 25 years) who were registered with a GP within QResearch Database between 1^st^ January 1998 and 31^st^ December 2019.

Cohort entry will be the latest of date of registration with the practice plus 1 year, date on which the practice computer system was installed plus 1 year, and the study start date (1 January 1998) and for those who have cancer-relevant clinical features the first date in which the clinical feature was recorded. Exit from the cohort will be the earliest of 6 months following study entry date, 6 months following first recorded cancer-relevant symptom, or cancer diagnosis. Analyses will be restricted to CTYA who had a cancer diagnosis within 6 months or CTYA who had at least 6 months follow-up.

Cases will be defined as the commonest non-skin cancer diagnoses in this age group^23^ and will be categorised into subtypes according to the International Classification for Childhood Cancers (third edition, ICCC-3)^24^: 1) Leukaemias and myelodysplastic diseases, 2) lymphomas and reticuloendothelial neoplasms, 3) central nervous system and intraspinal tumours, 4) soft tissue and bone sarcomas, 5) abdominal tumours (renal tumours, neuroblastomas, hepatoblastomas) and 6) germ cell, trophoblastic and other gonadal tumours. Cases and their date of diagnosis will be identified through the National Cancer Registry. Cases with a diagnosis prior to study start date were excluded, as were those with the following pre-existing conditions linked to cancer: Down’s Syndrome, neurofibromatosis type I and II, ataxia telangiectasia, tuberous sclerosis, and Li Fraumeni ^25-29^. Incidence rates will be calculated for childhood (0-14 years) and TYA (15-24 years) and compared to available national incidence rates.

#### Outcome of Interest

The outcome of interest will be a diagnosis of the selected CTYA cancers within/at 6 months from presentation:

1. CTYA blood cancers (0-24 years): Any leukaemia/lymphoma diagnosis
2. CTYA solid cancers (0-24 years): Any CNS/sarcoma/abdominal tumour/gonadal germinal tumour diagnosis

Any CTYA who has a cancer diagnosis prior to cohort entry or after cohort exit will be excluded.

#### Selection of Clinical Features and Risk Factors (Candidate Predictor Variables)

Table 1 details all identified candidate predictor variables for model development. Cancer-associated clinical features in CTYA will be selected through previously published evidence available^30-33^. These clinical features (includes symptoms, signs and blood test results) will be identified through QResearch Database and results determined to be incorrect/outliers by the clinical team will be excluded. All blood test results will be categorised into normal and abnormal according to nationally available laboratory cut-off values^34^. Any clinical feature before cohort entry or after cohort exit will be excluded.

**Table 1.** Candidate Predictor Variables for Model development.

|  | ***Variables*** |
| --- | --- |
| Demographic | Age |
|  | Sex |
|  | Deprivation Level |
|  | Ethnicity |
| Clinical Features | Organomegaly/abdominal mass |
|  | Lymphadenopathy |
|  | Fever |
|  | Limb pain |
|  | Joint pain |
|  | Limp/abnormal gait |
|  | Bruising |
|  | Tiredness |
|  | Looks anaemic/pale |
|  | Abdominal pain |
|  | Rash |
|  | Abnormal skin lesions |
|  | Cough |
|  | Chest pain |
|  | Headache |
|  | Vomiting |
|  | Head/neck lump |
|  | Lump on body |
|  | Hemiparesis |
|  | Squint |
|  | Visual acuity problems |
|  | Seizures |
|  | Haematuria |
|  | Feels unwell |
|  | Constipation |
|  | Dizziness |
| Blood Tests | Haemoglobin |
|  | White cell count (inc. differential count) |
|  | Platelet count |
|  | Mean Cell Volume |
|  | C-reactive protein |
|  | Erythrocyte sedimentation rate |
|  | Ferritin |
|  | Lactate Dehydrogenase |
|  | Alanine transaminase (ALT) |
|  | Bilirubin |
|  | Albumin |

Record of sociodemographic risk factors on the study entry date will be used for data extraction. Ethnicity will be defined as self- or parent-reported ethnicity on primary care health records. Ethnic groups are recorded based on the 2011 Census of England and Wales in 2 broad categories (White, Other)^35^. Level of deprivation will be assessed through the Townsend deprivation score which is an area-level continuous score based on an individual’s postcode; factors that included unemployment, non–car ownership, non–home ownership, and household overcrowding, are measured for a given area of approximately 120 households, via the 2011 Census of England and Wales and combined to give a Townsend score for that area, with the first quintile representing the lowest deprivation level and the fifth quintile representing the highest deprivation level^36^.

#### Sample Size Calculations

Sample size calculations were carried out using ‘pmsampsize’ on Stata^37^. We set our time point at 6 months and used 15% of the maximum permitted Cox-Snell R-squared (derived from Riley et al., 2020^37^) as there are no previous risk prediction models for its derivation. Cancer Research UK data were used to estimate incidence of cancers. We provide sample size calculations for the following cohorts: 1) blood cancers (leukaemias and lymphomas), 2) solid cancers (CNS, renal and hepatic tumours, soft tissue and bone sarcomas and neuroblastomas) (Table 2).

**Table 2.** Sample size requirements for clinical prediction model development using Riley et al., 2020^37^.

| Cohort | $R^2_{cs}$ | Parameters | Diagnosis Rate | Required Sample size | Events per predictor parameter |
| --- | --- | --- | --- | --- | --- |
| Blood cancers (0-24 years) | 0.00027 | 30 | 0.000089 (89 per million) | 999850 | 3 |
| Solid Tumours (0-24 years) | 0.0003 | 30 | 0.0001 (100 per million) | 870818 | 3 |

Previous studies using QResearch have identified a cohort of ∼5 million children and young adults^38^, with approximately 2 million children and 3 million teenagers and young adults. Altogether, this indicates that our study has sufficient sample size.

#### Model Derivation

We will explore two modelling approaches for model derivation:

1. Cox proportional hazards model
2. Logistic regression model

First, a full model will be fitted using all candidate predictors in all imputed datasets and combined using Rubin’s rules. Second, the pooled model will be used to select any categorical predictor with exponentiated coefficients >1.1 or <0.9 (at p<0.01) and any continuous predictors with significance of p<0.01. Third, these selected predictors will be used to refit the final model. This is to ensure both clinical and statistical magnitude of predictors are considered. Interactions between variables will be considered (based on a clinical plausibility) and interaction terms included within the model development process. Model coefficients will be combined, and in the case of Cox models with the baseline survival function, in order to calculate the linear predictor. Finally, model performance will be assessed through calculating the apparent discrimination, using Harrell’s C-index if cox regression and area-under-the-curve if logistic regression, and calibration, using calibration-in-the-large, calibration slope and smoothed calibration plots.

For cox regression models, proportional hazard assumptions will be assessed using Schoenfeld residuals.

#### Model Validation

The model derivation process will be validated using the internal-external cross validation approach^39^. This approach allows for the assessment of overall model performance as well as potential transportability of the model to a ‘distinct’ population and takes advantage of the availability of data from different geographical locations within QResearch Database (up to 10 locations across England). The summary of this approach is as follows: first, one geographical region will be ‘excluded’ whilst the model is developed using all other available regions. Second, data from the ‘excluded’ geographical region will be used to assess model performance using the aforementioned performance measures. These two steps will be repeated for each geographical region. Finally, random effects meta-analysis using the Hartung-Knapp-Sidik-Jonkmann method^40^ will be carried out to pool region-level performance measures and provide a pooled estimate of performance measures (i.e., discrimination and calibration).

#### Missing Data and other Statistical Consideration

We anticipate that there will be missing data for deprivation level (Townsend Quintile), ethnicity and blood test values and we consider these variables under the missing at random assumption^41^. For categorical variables we will use multinomial logistic regression, for ordinal variables ordinal logistic regression for imputation. 5 imputations will be carried out to strike a balance between % missingness and computational efficiency. Model coefficients will be pooled in accordance with Rubin’s rules. The imputation model will be inclusive of candidate variables and the outcome variable.

Any continuous candidate variable (e.g., age) will be assessed for nonlinearity and handled using fractional polynomials. Fractional polynomials will be fitted prior to imputation analyses. Clustering of participants within individual general practices will be accounted for using clustered standard errors.

#### Decision Curve Analysis

To assess potential clinical utility, a decision curve analysis will be used to compare standard net clinical benefit (i.e., the trade-off between the benefits of true positives and harms of false positives) using developed models with a scenario where no model is used.

#### Statistical Software

All analyses will be carried out using Stata (v17)^42^.

#### Patient and Public Involvement

National charities were approached to identify patient representatives and currently two representative young people have volunteered to provide input in study design, interpretation and dissemination of results.

#### Ethics and Dissemination

This project (OX94) has been approved by the QResearch scientific committee. The QResearch database annually obtains ethical approval from the East Midlands-Derby Research Ethics Committee (REC reference 18/EM/0400).

## Data Availability

N/A

## Notes

### Competing Interest Statement

JHC reports grants from National Institute for Health Research Biomedical Research Centre, Oxford, grants from John Fell Oxford University Press Research Fund, grants from Cancer Research UK (CR-UK) grant number C5255/A18085, through the Cancer Research UK Oxford Centre, grants from the Oxford Wellcome Institutional Strategic Support Fund (204826/Z/16/Z), during the conduct of the study. JHC is an unpaid director of QResearch, a not-for-profit organisation which is a partnership between the University of Oxford and EMIS Health who supply the QResearch database used for this work. Until 09 Aug 2023, JHC had a 50% shareholding in ClinRisk Ltd, co-owning it with her husband, who was an executive director. On 9th August 2023, 100% of the share capital was donated to Endeavour Health Care Charitable Trust and the company renamed to Endeavour Predict Ltd. JHC is an unpaid consultant to Endeavour Predict Ltd and her husband is a non-executive director to cover the transition. The company licences software both to the private sector and to NHS bodies or bodies that provide services to the NHS (through GP electronic health record providers, pharmacies, hospital providers and other NHS providers). This software implements algorithms (including QCancer) developed from access to the QResearch database during her time at the University of Nottingham.

### Funding Statement

Cancer Research UK EDDCPJT\100016. The funding body had no role in the design and conduct of the study; collection, management, analysis, and interpretation of the data; preparation, review, or approval of the manuscript; and decision to submit the manuscript for publication.

### Author Declarations

This project (OX94) has been approved by the QResearch scientific committee at the University of Oxford. The QResearch Database annually obtains ethical approval from the East Midlands-Derby Research Ethics Committee (REC reference 18/EM/0400).

